# Aetiological differences between novel subtypes of diabetes derived from genetic associations

**DOI:** 10.1101/2020.09.29.20203935

**Authors:** Dina Mansour Aly, Om Prakash Dwivedi, Rashmi B Prasad, Annemari Käräjämäki, Rebecka Hjort, Mikael Åkerlund, Anubha Mahajan, Miriam S. Udler, Jose C Florez, Mark I McCarthy, Regeneron Genetics Center, Julia Brosnan, Olle Melander, Sofia Carlsson, Ola Hansson, Tiinamaija Tuomi, Leif Groop, Emma Ahlqvist

## Abstract

**Background:** Type 2 diabetes (T2D) is a multi-organ disease defined by hyperglycemia resulting from different disease mechanisms. Using clinical parameters measured at diagnosis (age, BMI, HbA1c, HOMA2-B, HOMA2-IR and GAD autoantibodies) adult patients with diabetes have been reproducibly clustered into five subtypes, that differed clinically with respect to disease progression and outcomes.^1^ In this study we use genetic information to investigate if these subtypes have distinct underlying genetic drivers.

**Methods:** Genome-wide association (GWAS) and genetic risk score (GRS) analysis was performed in Swedish (N=12230) and Finnish (N=4631) cohorts. Family history was recorded by questionnaires.

**Results:** Severe insulin-deficient diabetes (SIDD) and mild obesity-related diabetes (MOD) groups had the strongest family history of T2D. A GRS including known T2D loci was strongly associated with SIDD (OR per 1 SD increment [95% CI]=1.959 [1.814-2.118]), MOD (OR 1.726 [1.607-1.855]) and mild age-related diabetes (MARD) (OR 1.771 [1.671-1.879]), whereas it was less strongly associated with severe insulin-resistant diabetes (SIRD, OR 1.244 [1.157-1.337]), which was similar to severe autoimmune diabetes (SAID, OR 1.282 [1.160-1.418]). SAID showed strong association with the GRS for T1D, whereas the non-autoimmune subtype SIDD was most strongly associated with the GRS for insulin secretion rate (*P*<7.43×10^−9^). SIRD showed no association with variants in *TCF7L2* or any GRS reflecting insulin secretion. Instead, only SIRD was associated with GRS for fasting insulin (*P*=3.10×10^−8^). Finally, a T2D locus, rs10824307 near the *ZNF503* gene was uniquely associated with MOD (OR_meta_=1.266 (1.170-1.369), *P*=4.3×10^−9^).

**Conclusions:** New diabetes subtypes have partially different genetic backgrounds and subtype-specific risk loci can be identified. Especially the SIRD subtype stands out by having lower heritability and less involvement of beta-cell related pathways in its pathogenesis.

**Research in context:** *Evidence before this study:* In March 2018 we suggested a novel subclassification of diabetes into five subtypes. This classification was based on clustering using clinical parameters commonly measured at diabetes diagnosis (age at diabetes onset, HbA1c, bodymass index, presence of GAD autoantibodies and HOMA2 indices for insulin resistance and secretion). These subtypes differed with respect to clinical characteristics, disease progression and risk of complications, but it remained unclear to what extent these subtypes have different underlying pathologies. In our original publication we analysed a small set of genetic risk variants for diabetes and found differential associations between subtypes, suggesting potential aetiological differences.

*Added value of this study:* In this study we have conducted a full genome analysis of the original ANDIS cohort, including genome-wide association studies and polygenic risk score analysis with replication in an independent cohort. We have also compared heritability and prevalence of having a family history of diabetes in the subtypes.

*Implications of all the available evidence:* We demonstrate that stratification into subtypes facilitates identification of genetic risk loci and that the aetiology of the subtypes is at least partially distinct. These results are especially important for the future study and treatment of individuals belonging to the severe insulin-resistant diabetes (SIRD) subtype, whose pathogenesis appears to differ substantially from that of traditional T2D.

## Introduction

Although defined by a single metabolite, glucose, T2D is increasingly recognized as a highly heterogeneous disease, including individuals with varying clinical characteristics, disease progression, drug response and risk of complications.^1^ Traditionally T2D is thought to develop when a person can no longer increase insulin secretion to compensate for the increased demand imposed by insulin resistance, which could be due to a number of pathways. Classification into subtypes with different risk profiles and disease aetiologies at diagnosis could enable tailored treatment to address the main dysfunction in each patient, and resources could be focused on those most likely to develop complications.

To identify more homogeneous patient groups we recently performed a cluster analysis in a large Swedish cohort of patients with diabetes (ANDIS, All New Diabetics In Scania), based on six clinical variables: GAD autoantibodies (GADA), body mass index (BMI), glycated hemoglobin (HbA1c), age at diabetes diagnosis, insulin secretion estimated as HOMA2-B and insulin sensitivity as HOMA2-IR derived from fasting glucose and C-peptide.^1^ We identified five clusters of patients with different clinical characteristics, disease progression and outcomes, and suggested the following new sub-classification of adult diabetes. Severe Autoimmune Diabetes (SAID, 6% of patients), is defined by presence of GADA and includes patients traditionally referred to as type 1 diabetes (T1D) and Latent Autoimmune Diabetes in Adults (LADA). This subtype is characterized by early onset, low insulin secretion, relatively low BMI and poor metabolic control.^1^ Severe Insulin-Deficient Diabetes (SIDD, 18%), is, despite GADA negativity, also characterized by early onset, low insulin secretion, relatively low BMI and poor metabolic control. SIDD also has a high risk of developing diabetic retinopathy^1^ and neuropathy.^2^ Severe Insulin-Resistant Diabetes (SIRD, 15%), is characterized by late onset, obesity, insulin resistance and high risk of diabetic nephropathy and non-alcoholic fatty liver disease (NAFLD).^1,2^ Moderate Obesity-related Diabetes (MOD, 22%) has early onset, obesity and a relatively mild disease in terms of progression of hyperglycaemia and complications. Moderate Age-Related Diabetes (MARD, 39%), is characterized by late onset diabetes and relatively good metabolic control. MOD and MARD have the lowest risk of diabetic complications.^1,2^ Subsequent studies have replicated these five clusters in other cohorts and further showed that the prevalence of diabetic complications and treatment response differs between clusters, exemplified by a high frequency of fatty liver disease and diabetic nephropathy as well as a better response to insulin sensitizers in the SIRD group, and increased prevalence of neuropathy in the SIDD group.^2-4^

The aim of this study was to use genetic information to elucidate to what extent these clusters represent aetiologically distinct subtypes of diabetes with different genetic risk profiles, by performing global (genome-wide) and restricted genetic risk score (GRS) analysis in the clusters. We also hypothesized that sub-classification of T2D would allow identification of new loci affecting specific disease pathways.

## Results

### Heritability of diabetes subtypes

The heritability of the subtypes was compared using two strategies: presence of self-reported family history of diabetes (FHD) and SNP-wide heritability based on GWAS in ANDIS, searching for association with subtypes: SAID (N=452), SIDD (N=1193), SIRD (N=1130), MOD (N=1374) and MARD (N=2861), and using 2744 individuals without diabetes from the MDC cohort as controls (Table S1).

Family history of diabetes differed between subtypes (Figure 1, Table S2). Having a first or second degree relative with T2D was most common in the MOD (87.8%) and SIDD (83.7%) subtypes, and least common in SAID (72.8%). Of the GADA negative subtypes, MARD and SIRD had the least family history of T2D (76.8 and 77.6% respectively). As expected, SAID had the highest frequency of first- and second-degree relatives with T1D (7.7%), defined as age at diagnosis before 40 years and insulin-treatment.

**Figure 1.**
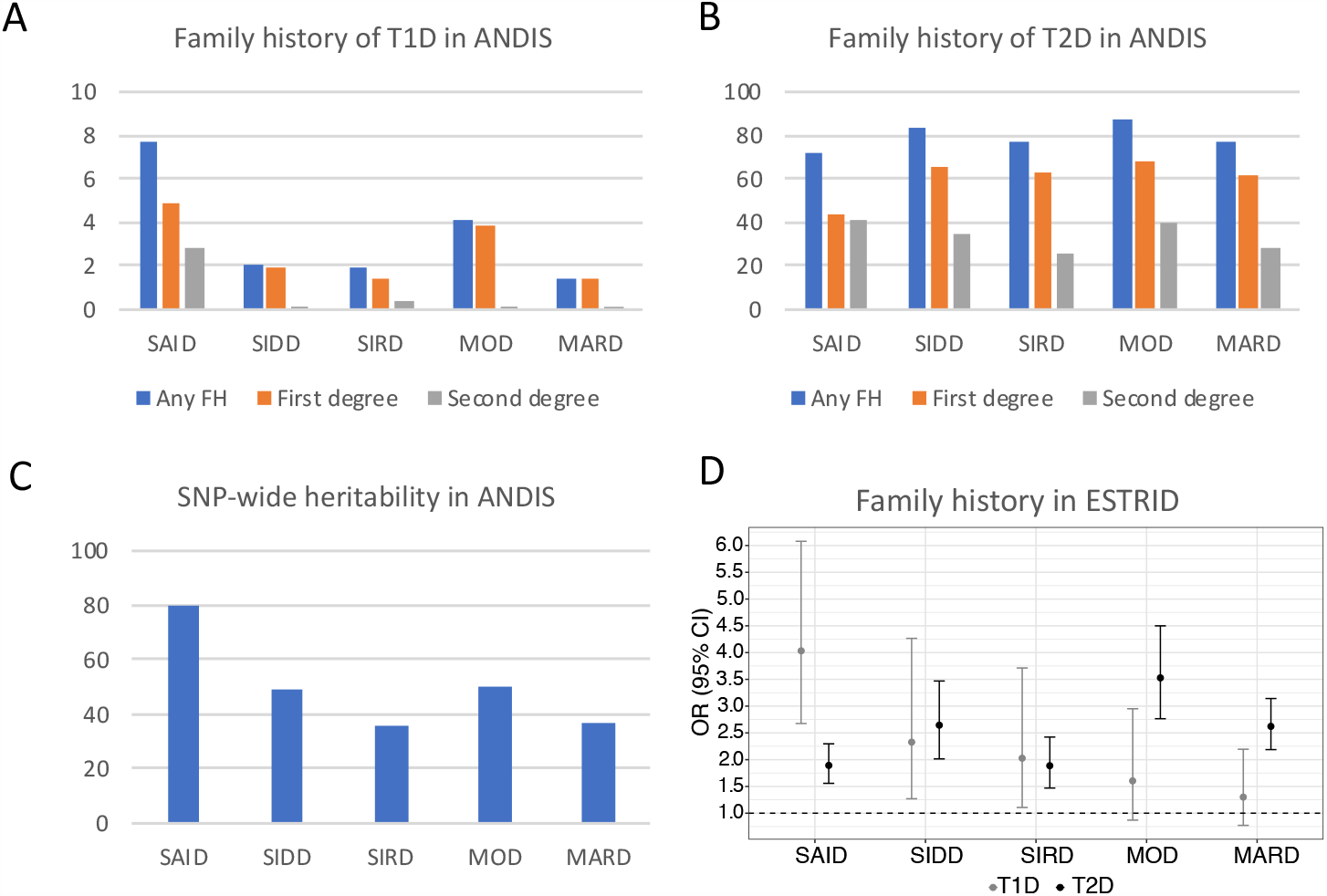
Total heritability and family history of diabetes differs between subtypes. Percent of participants with any family history (blue), first (orange) and second (grey) degree family history of diabetes. A) Family history of T1D in ANDIS, defined by insulin treatment and onset before age 40 differed between all subtypes (p=1.72⨯10^−11^) and between GADA negative subtypes (p=1.2⨯10^−5^). B) Family history of T2D in ANDIS defined as all other diabetes (onset after 40 years or no insulin treatment) also differed between all subtypes (p=3.2⨯10^−17^) and between GADA negative subtypes (p=2.7⨯10^−15^). C) Heritability, estimated as genetic variance/phenotypic variance ratio, based on all genotyped SNPs. D) Risk associated with family history of T1D and T2D in ESTRID (a sub-study of ANDIS) for each subtype compared to non-diabetic controls. Exact numbers and statistics in tables S2-S5.

Analysis of a subset of ANDIS participating in the ESTRID study,^5^ showed increased risk associated with family history of T2D for all subtypes compared to matched diabetes-free controls, with the lowest risk in SIRD for having first degree (OR=2.680 [2.060-3.487] and second degree (OR=0.753 [0.517-1.096]), Table S3-4) T2D relatives.

SNP-wide heritability was calculated using the Genome-wide complex trait analysis (GCTA) Genome-based restricted maximum likelihood (GREML) method,^6^ as genetic variance/phenotypic variance ratio (VG/VP[SE]). Of the GADA negative subtypes, SIDD and MOD had the highest heritability (0.494 [0.047] and 0.500 [0.042], respectively), and SIRD and MARD the lowest (0.354 [0.045] and 0.364 [0.031] (Figure 1C, Table S5).

### Genetic risk score analysis for T1D and T2D

We first constructed a global polygenic T2D risk score (T2D-gPRS), including all genotyped SNPs (n=389,243) weighted by their effect on T2D risk in the European DIAMANTE study.^7^ In line with the FHD data, the T2D-gPRS showed increased genetic risk for all diabetes subtypes with the highest risk for SIDD and MOD, and lower risk for SAID, SIRD and MARD (Figure 2A, Table S6). The quintile with the highest T2D-gPRS had 5.927 (4.627-7.630) times higher risk of SIDD and 8.712 (6.816-11.216) times higher risk of MOD compared to the lowest quintile, but only 3.383 (2.651-4.326) times higher risk of SIRD and 3.213 (2.670-3.873) times higher risk of MARD (Figure 2A right panel, Table S6).

**Figure 2.**
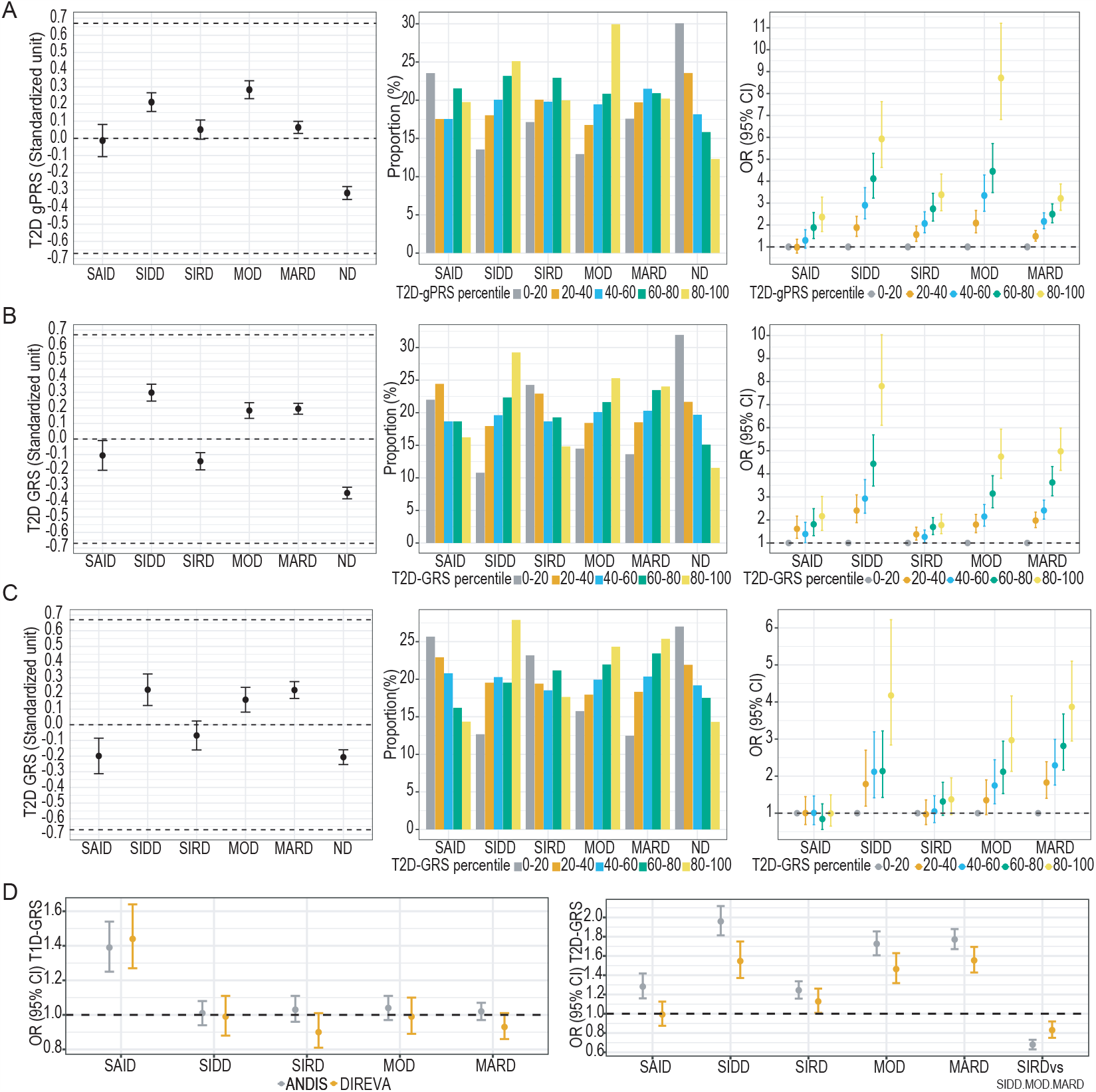
Distribution and association of T1D and T2D genetic risk scores with diabetes subtypes. A-C Left panels show mean T2D genetic risk scores (95% CI). Middle panels show proportions of individuals in T2D risk score quintiles for each group. Right panels show risk (OR, 95%CI) of each diabetes subtype per increasing T2D genetic risk score quintile compared to non-diabetic controls (ND) using the lowest genetic risk score quintile (0-20) as a reference. A) Global T2D polygenic risk score (T2D-gPRS) distribution in ANDIS. The T2D-gPRS was constructed using all the genome-wide SNPs (N=389,243) after clumping weighted by effect size from the largest European T2D meta-analysis study [1]. B) T2D restricted genetic risk score (T2D-GRS) distribution in ANDIS. The T2D-GRS was calculated by including only genome wide significant variants (384 out of 403 T2D risk score loci from Mahajan et al, Table S7). C) T2D-GRS (as described in B) in DIREVA. D) Association of T1D genetic risk score (T1D-GRS), calculated using all variants (n=66, Table S7) associated with T1D at genome wide significant levels in the largest European T1D fine-mapping study, and T2D-GRS (as in B) with diabetes subtypes in ANDIS and DIREVA. Association analysis was done using logistic regression compared to non-diabetic controls and/or other groups as specified. Exact numbers and statistics in Table S6.

Second, we generated restricted T2D (n=384, T2D-GRS) and T1D GRS (n=66 non-HLA SNPs, T1D-GRS) based on previously reported genome-wide significant SNPs (Table S7). In ANDIS, the T2D-GRS showed the highest genetic risk for SIDD (OR per 1 SD increment [95% CI]=1.959 [1.814-2.118]), MOD (OR 1.726 [1.607-1.855]) and MARD (OR 1.771 [1.671-1.879]), with significantly lower risk for SIRD (OR 1.244 [1.157-1.337], *P*_SIRDvsSIDD+MOD+MARD_=6.71×10^−29^) and SAID (OR 1.282 [1.160-1.418], Figure 2D, Table S6). Here, the quintile with the highest T2D-GRS had an OR of only 1.777 (1.402-2.250) in SIRD but 7.804 (6.107-10.032) in SIDD (Figure 2B right panel, Table S6). The same pattern was seen in the Finnish DIREVA cohort (Figure 2C, Table S6). The T1D-GRS, showed association exclusively with the autoimmune SAID group (*P*≤ 3.36×10^−8^, Figure 2D, Table S6).

### Weighted T2D genetic risk score analysis for insulin secretion and sensitivity measures

We then constructed weighted genetic risk scores (T2D-wGRS) using known T2D associated variants^7^ weighted by their genetic effect on measures of insulin secretion and sensitivity, extracted from their respective largest genetic studies (n=219 to 319 with data available, Table S7 and S8, Figure 3). T2D-wGRS for first phase insulin response, measured either as insulin secretion rate (ISR-wGRS) based on serum C-peptide during an intraveneous glucose tolerance test (IVGTT),^8^ or corrected insulin response (CIR-wGRS) during an oral glucose tolerance test (OGTT), ^9^ had the strongest association with SIDD (*P*≤5.03×10^−7^) but no association with SIRD (*P*>0.5, Figure 3A and B, Table S8) in ANDIS. In contrast, T2D-wGRS for fasting insulin (F-INS-wGRS)^10^ was only associated with SIRD (*P*=3.10×10^−8^) with no significant association with other subtypes (*P*>0.1). A similar pattern of association was observed in DIREVA for SIRD, with association with F-INS-wGRS (*P*=1.70×10^−5^) and no association with insulin secretion wGRS (Figure 3B, Table S8). An insulin sensitivity index score (ISI-wGRS)^11^ was most strongly associated with SIRD (*P*=2.93×10^−6^) and MOD (*P*= 1.87×10^−4^) but showed nominal or no association with MARD (*P*=0.064) and SIDD (*P*=0.029) in ANDIS.

**Figure 3.**
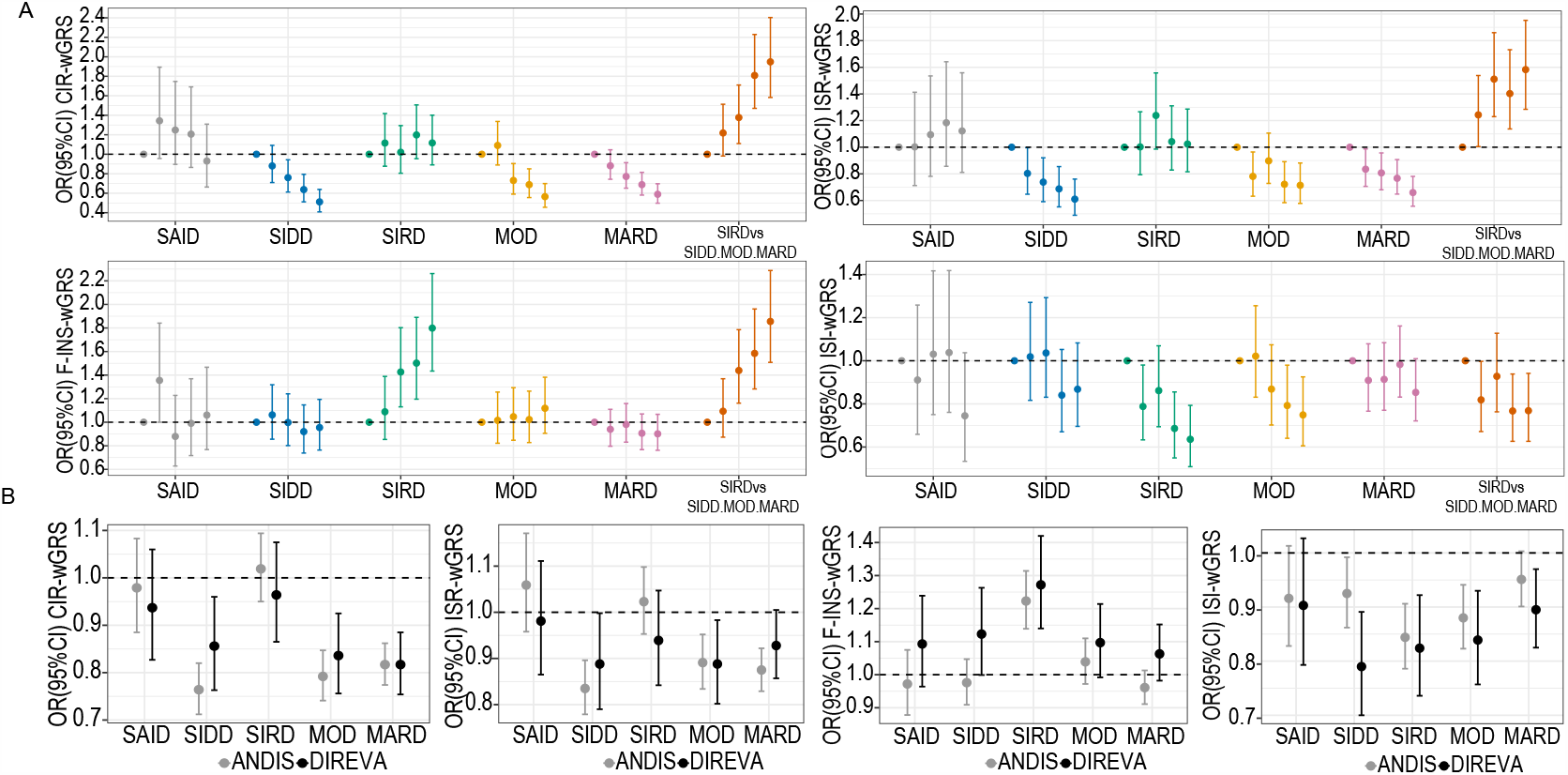
Association of insulin secretion and insulin sensitivity GRS with diabetes subtypes. T2D weighted GRS (T2D-wGRS) for diabetes subphenotype traits were calculated using known T2D associated SNPs (n=219 to 319, Table S7) weighted by effect sizes obtained from the largest meta-analyses for measures of insulin secretion: Corrected insulin response at 30 min during OGTT (CIR-wGRS), C-peptide based insulin secretion rate during IVGTT (ISR-wGRS), fasting insulin (F-INS-wGRS) and Insulin sensitivity index during an OGTT (ISI-wGRS). A) Association of increasing quintiles of each T2D-wGRS compared to non-diabetic controls, and/or other groups as specified, using lowest quintile as reference in ANDIS-MDC. B) Association of each T2D-wGRS with subtype compared to non-diabetic controls in ANDIS-MDC and DIREVA-Botnia. Association analysis was done using logistic regression. Exact numbers and statistics in Table S8.

Fasting and two-hour glucose wGRSs showed strong associations with SIDD, MOD and MARD (*P*≤9.70×10^−15^) and weaker associations (*P*≥6.15×10^−4^) with SAID and SIRD (Table S8). A wGRS for fasting proinsulin, showed nominal associations with SIDD (*P*=0.003) and MARD (*P*=0.021) (Table S8) in ANDIS.

### Genetic risk score analysis for lipids and weight related phenotypes

Further, we constructed GRS, based on previously reported genome-wide significant SNPs (*P*<5×10^−8^, n=39 to 518) for traits related to weight and blood lipids (Fig. 4, Table S7 and S9). A BMI-GRS was most strongly associated with MOD (OR=1.290 [1.206-1.379], *P*=9.80×10^−14^), whereas MARD showed no association (*P*=0.099). A similar pattern of associations was observed for GRS of waist circumference and predicted visceral adiposity (VAT) in ANDIS (Fig.4 and Table S9). A GRS for waist/hip ratio (WHR-GRS) was associated with both SIRD (*P*=2.89×10^−4^) and MOD (*P*=3.84×10^−3^). However, a GRS for BMI adjusted WHR showed the strongest association with SIRD (*P*=7.59×10^−5^) but no association with MOD (Fig.4 and Table S9). GRS for triglycerides showed no association but GRS for cholesterol levels (HDL, LDL, total cholesterol) were associated with SIRD with nominal significance (*P*<0.02) in ANDIS (Fig 4, Table S9).

**Figure 4.**
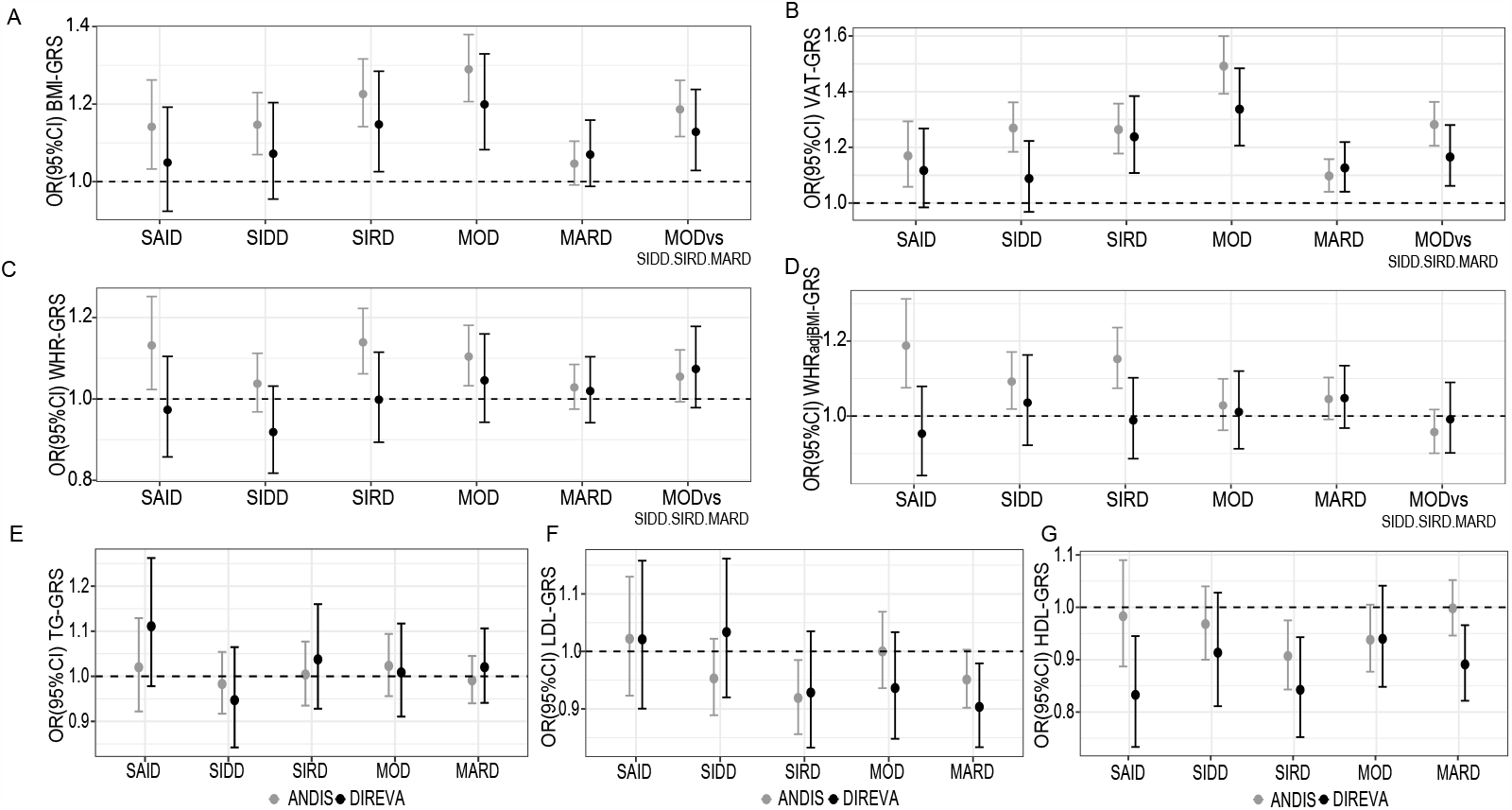
Association of genetic risk scores of adiposity related traits and lipids with diabetes subtypes. Association of genetic risk scores for A) body mass index (BMI), B) predicted visceral adipose tissue (VAT), C) waist-hip ratio (WHR), D) WHR adjusted for BMI, E-G) lipids traits (TG-triglycerides, HDL-high density lipoprotein, LDL-low density lipoprotein). Genetic risk scores were constructed using only genome wide significant (p<5×10^−8^, n=39 to 518) SNPs retained after clumping for each trait using their respective largest published meta-analysis summary data (Tables S7, Method). Association analysis was done using logistic regression compared to non-diabetic controls and/or other groups as specified. Exact numbers and statistics in Table S9.

Finally, we tested published GRS based on clustering of T2D risk variants by Mahajan *et al*. (Table 2 and S10)^12^. Three independent GRSs for insulin secretion were associated with SIDD, MOD and MARD (*P*≤5.84×10^−7^) but none was associated with SIRD (*P*>0.2). The insulin action GRS was associated with all T2D-clusters (*P*≤7.44×10^−5^). The GRS for impaired lipids showed some association with SIRD (OR=1.09 [1.01-1.17], *P*=0.024) but not with MOD (*P*>0.9). The adiposity risk score was most strongly associated with MOD (OR=1.2 [1.12-1.28], *P*=1.87×10^−7^) and the least with MARD (OR=1.06 [1.00-1.10], *P*=0.039).

**Table 1.** Genome-wide significant loci in ANDIS with replication in DIREVA.

|  | ANDIS |  | DIREVA |  | Meta-analysis |  |
| --- | --- | --- | --- | --- | --- | --- |
| Cluster | OR(95% CI) | P | OR(95% CI) | P | OR(95% CI) | P |
| **HLA-DQB1 rs9273368** |  |  |  |  |  |  |
| SAID | 2.678 (2.314-3.100) | $6.5 \times 10^{-40}$ | 2.893(2.448-3.418) | $1.14 \times 10^{-35}$ | 2.769(2.481-3.091) | $1.34 \times 10^{-73}$ |
| SIDD | 0.877 (0.786-0.979) | 0.018 | 1.411(1.208-1.648) | $1.37 \times 10^{-5}$ | 1.027(0.939-1.123) | 0.559 |
| SIRD | 0.984(0.883-1.097) | 0.750 | 1.033(0.887-1.203) | 0.679 | 1.000(0.916-1.092) | 0.997 |
| MOD | 1.027(0.927-1.138) | 0.586 | 1.019(0.891-1.203) | 0.78 | 1.024(0.944-1.111) | 0.567 |
| MARD | 0.998(0.918-1.0849) | 0.915 | 0.975(0.880-1.081) | 0.635 | 0.989(0.927-1.055) | 0.732 |
| **TCF7L2 rs7903146** |  |  |  |  |  |  |
| SAID | 1.103(0.937-1.299) | 0.235 | 0.970(0.784-1.200) | 0.779 | 1.052(0.924-1.197) | 0.445 |
| SIDD | 1.559(1.394-1.745) | $8.6 \times 10^{-15}$ | 1.252(1.045-1.501) | 0.015 | 1.467(1.334-1.614) | $3.0 \times 10^{-15}$ |
| SIRD | 1.072(0.955-1.204) | 0.230 | 1.127(0.945-1.345) | 0.183 | 1.088(0.988-1.199) | 0.086 |
| MOD | 1.402(1.262-1.557) | $3.1 \times 10^{-10}$ | 1.236(1.061-1.439) | 0.00635 | 1.346(1.235-1.468) | $1.8 \times 10^{-11}$ |
| MARD | 1.417(1.302-1.542) | $6.1 \times 10^{-16}$ | 1.543(1.377-1.728) | $6.35 \times 10^{-14}$ | 1.461(1.365-1.563) | $8.6 \times 10^{-28}$ |
| **ZNF503 rs10824307** |  |  |  |  |  |  |
| SAID | 1.033(0.886-1.203) | 0.653 | 0.936(0.788-1.111) | 0.449 | 0.989(0.882-1.109) | 0.848 |
| SIDD | 1.077(0.968-1.198) | 0.177 | 0.995(0.851-1.163) | 0.949 | 1.050(0.962-1.147) | 0.276 |
| SIRD | 1.133(1.018-1.262) | 0.014 | 1.056(0.913-1.221) | 0.465 | 1.105(1.014-1.205) | 0.023 |
| MOD | 1.347(1.219-1.487) | $1.3 \times 10^{-9}$ | 1.144(1.007-1.301) | 0.040 | 1.266(1.170-1.369) | $4.3 \times 10^{-9}$ |
| MARD | 1.104(1.017-1.198) | 0.007 | 1.031(0.934-1.138) | 0.547 | 1.074(1.008-1.144) | 0.028 |

**Table 2.** GRS based on SNPs clustered by function from Mahajan et al.

|  | SAID |  | SIDD |  | SIRD |  | MOD |  | MARD |  |
| --- | --- | --- | --- | --- | --- | --- | --- | --- | --- | --- |
|  | OR (95%CI) | P | OR (95%CI) | P | OR (95%CI) | P | OR (95%CI) | P | OR (95%CI) | P |
| **Adiposity** | **1.11 (1.00-1.23)** | **0.047** | **1.10 (1.02-1.18)** | **0.009** | **1.11 (1.03-1.19)** | **0.004** | **1.20 (1.12-1.28)** | **1.87x10<sup>-7</sup>** | **1.06 (1.00-1.12)** | **0.039** |
| **Insulin action** | 1.08 (0.98-1.20) | 0.126 | **1.17 (1.09-1.25)** | **1.52x10<sup>-5</sup>** | **1.17 (1.09-1.26)** | **9.75x10<sup>-6</sup>** | **1.14 (1.07-1.22)** | **7.44x10<sup>-5</sup>** | **1.16 (1.10-1.23)** | **4.03x10<sup>-8</sup>** |
| **Insulin action/secretion** | **1.24 (1.12-1.37)** | **3.87x10<sup>-5</sup>** | **1.23 (1.15-1.32)** | **7.43x10<sup>-9</sup>** | 1.04 (0.97-1.11) | 0.307 | **1.19 (1.12-1.28)** | **1.90x10<sup>-7</sup>** | **1.19 (1.13-1.26)** | **2.16x10<sup>-10</sup>** |
| **Insulin secretion 1** | 1.01 (0.91-1.12) | 0.874 | **1.31 (1.22-1.41)** | **1.37x10<sup>-13</sup>** | 1.04 (0.97-1.12) | 0.242 | **1.26 (1.18-1.35)** | **3.39x10<sup>-11</sup>** | **1.29 (1.22-1.37)** | **3.07x10<sup>-20</sup>** |
| **Insulin secretion 2** | 0.99 (0.90-1.10) | 0.894 | **1.26 (1.18-1.36)** | **4.18x10<sup>-11</sup>** | 1.02 (0.95-1.09) | 0.665 | **1.18 (1.11-1.27)** | **5.84x10<sup>-7</sup>** | **1.23 (1.17-1.30)** | **3.85x10<sup>-14</sup>** |
| **Impaired lipids** | 1.08 (0.97-1.20) | 0.167 | 1.07 (1.00-1.15) | 0.056 | **1.09 (1.01-1.17)** | **0.025** | 1.00 (0.93-1.07) | 0.987 | **1.06 (1.00-1.12)** | **0.033** |

We also compared the association with GRS developed by Udler *et al* ^13^ (Table S11). Again, GRS for beta-cell function was most strongly associated with SIDD (OR=1.32 [1.23-1.42], *P*=1.1×10^−14^) but not with SIRD (OR=1.01 [0.94-1.08], *P*>0.8). The proinsulin cluster was most strongly associated with SIDD (OR=1.14 [1.06-1.22], *P*=2.63×10^−4^) but also increased risk of MOD and MARD. In contrast, it was protective for SIRD (OR=0.93 [0.87-1.0], *P*=0.044).

### Genome-wide association analysis (GWAS)

Four loci reached genome-wide significance (*P*<5×10^−8^) in at least one subtype in ANDIS using diabetes-free individuals as controls (Table 1, Table S12). SAID was positively associated with variants in the *HLA-DQB1* & *HLA-DQA1* loci^14^ (OR_meta_=2.769 [2.481-3.091], *P*=1.34×10^−73^) whereas other clusters were not (*P*>0.5). The rs7903146 variant in the *TCF7L2* locus, previously shown as the variant most strongly associated with T2D and insulin secretion^15^ was associated with SIDD (OR_meta_=1.467 [1.334-1.614], *P*=3.0×10^−15^), MOD (OR_meta_=1.346 [1.235-1.468], *P*=1.8×10^−11^) and MARD (OR_meta_=1.461 [1.365-1.563], *P*=8.6×10^−28^). In contrast, there was no association between SIRD and the *TCF7L2* variant in either ANDIS or DIREVA (*P*>0.05).

The minor G allele of rs10824307 near the *ZNF503* gene was associated with increased risk of MOD (OR=1.372 [1.239-1519], *P*=1.31×10^−9^) with little association with other subtypes (*P*>0.01) in ANDIS. This association reached nominal significance (*P*=0.040) in the replication in DIREVA and remained significant in meta-analysis (OR_meta_=1.266 [1.170-1.369], *P*=1.39×10^−9^). A look-up of this SNP in the T2D Knowledge Portal (http://www.type2diabetesgenetics.org/) shows that it is associated with T2D at genome-wide significance in the AGEN+DIAMANTE study^16^ (OR=1.041, *P*=1.2×10^−8^), supporting that it is a true finding. Further, this SNP was associated with higher basal metabolic rate and higher whole-body fat-free mass (*P*=1.00×10^−11^) in the UK-biobank (*P*=2.50×10^−9^).^17,18^ Rs10824307 is an eQTL for the gene *C10ORF11* (*LRMDA*) in multiple tissues in the GTEx database,^19^ including adipose tissue (*P*=3.3×10^−42^) and pancreas (*P*=6.2×10^−18^), but is also associated with the expression of *ZNF503* (*P*=1.5×10^−7^) and *ZNF503-AS2* (*P*=1.4×10^−6^) in blood. All SNPs associated with at least one cluster at *P*<10^−5^ are shown in Table S12.

We also explored subtype association with previously published SNPs associated with T2D.^7^ Of 393 of these SNPs present in our GWAS, 66 were associated with at least one of the subtypes, and of them 5 with SAID, 28 with SIDD, 5 with SIRD, 19 with MOD and 34 with MARD (Table S13). In a GWAS of all GADA-negative individuals in the ANDIS dataset (N=9486) compared to diabetes-free MDC controls (N=2744) only 42 loci of the previous T2D-associated SNPs reached *P*<0.01 and 4 *P*<3.13×10^−5^ including *TCF7L2, TM6SF2, IGF2BP2*, and *ADCY5*.

## Discussion

The aim of this study was to leverage genome-wide genotyping data to investigate to what extent the newly defined subtypes of diabetes are genetically and thereby aetiologically distinct. The findings supported that the three severe subgroups, SAID, SIDD and SIRD have partially distinct pathogenesis, and that subclassification can improve power to detect diabetes loci, as shown for the MOD-specific *ZNF503* locus.

Heritability was strongest in MOD and SIDD, as shown by family history of diabetes, SNP-wide heritability estimates from genome-wide complex trait analysis (GCTA) and GRSs for T2D. Although a high burden of T2D SNPs increased the risk of all subtypes, this risk was only marginally higher for SIRD and MARD compared to the autoimmune subtype SAID. Although our results indicate that SIRD has a somewhat weaker genetic background compared to other subtypes, they also imply that the genetic component of this subtype has not been captured by GWAS that lump all patients with so-called T2D together. The relatively strong association with known loci compared to genome-wide SNPs for MARD, likely results from overrepresentation of this most common subtype in previous GWAS of T2D increasing power to identify loci associated with this subtype.

Though autoimmune SAID is mildly associated with T2D SNPs, it showed a strong and expected association with the HLA locus and the T1D GRS, which was not the case for the SIDD group, reinforcing the notion of its non-autoimmune origin. Further, SIDD showed clear association with variants in the classical T2D-associated gene, *TCF7L2*, as well as the strongest association with risk scores for insulin secretion. This is clinically relevant as the two subgroups have a similar presentation, with earlier onset, moderate overweight and similar degree of insulin deficiency, making it difficult for clinicians to differentiate between the two subgroups without antibody testing. By current care guidelines, SIDD is initially often treated with only metformin resulting in very poor metabolic control.^1^

Importantly, the insulin-resistant subtype, SIRD, showed no association with the *TCF7L2* locus or with any insulin secretion GRS, which is in line with the measured good insulin secretion. These results suggest that the beta cells play a different and minor role in the pathogenesis of SIRD. While the pancreas is often considered the most important organ in T2D, this might not be true for this subtype – at least regarding beta-cell function. Mirroring the high insulin-resistance index, the GRSs for insulin sensitivity showed the largest effect sizes for SIRD. Strikingly, SIRD was the only subtype associated with a GRS for fasting insulin (constructed based on association in non-diabetic individuals), which is considered to reflect liver insulin resistance. This goes well with the elevated liver lipids and higher risk scores for NAFLD observed in the SIRD subtype.^1,2^

These results clearly establish an aetiological difference between SIRD and MOD, two phenotypically close groups differentiated by the clustering approach. As expected, MOD has the highest GRS for BMI, WHR and VAT followed by SIRD, but in contrast to SIRD, MOD shows no association with fasting insulin GRS, or with the impaired lipids GRS suggesting a healthier type of obesity with less impact from liver insulin resistance. MOD was also associated with GRSs for insulin secretion. Thus, genetically, SIRD is characterized by association with insulin resistance whereas MOD represents a mix with both relative insulin deficiency and insulin resistance.

We have previously shown that the SIRD subtype develops diabetic kidney disease very early and that kidney function is often impaired already at diagnosis whereas SIDD develops early onset diabetic retinopathy.^1,2^ Identifying and treating these subtypes before they develop hyperglycemia could therefore be important to help prevent these complications.

The diagnostic accuracy of tests used to detect pre-diabetes in screening programs is low. HbA1c, FPG, and OGTT are used in the criteria for diabetes and prediabetes set by the American Diabetes Association and WHO, but the tests have shown different efficacy in their ability to detect prediabetes and results are highly variable depending on the population and setting^20^. The dissociation of genetic risk for increased fasting glucose, 2-hour glucose and fasting insulin suggests that different screening methods could be efficient at identifying the different diabetes subtypes.

Genetic risk scores for BMI, WHRadjBMI and ISI were not associated with MARD in ANDIS, indicating that obesity and insulin resistance may not be strong risk factors for this subtype. Instead, the MARD subtype was strongly associated with GRSs reflecting insulin secretion. This is in line with a stronger beneficial impact of sulfonylurea therapy, also suggesting a predominant role for insulin secretion defects in MARD.^3^

One of our original hypotheses was that we would be able to identify subtype specific loci that had escaped detection in previous GWAS. Indeed, we identified one such locus, the MOD-specific *ZNF503*/*LMDR*, providing a proof of principle for this idea. With an OR of approximately 1.3 in MOD patients, we reached genome-wide significance in our discovery analysis including only 1374 cases and 2744 controls. In the recent AGEN and DIAMANTE study^16^ including 433,530 individuals of East-Asian origin, the locus reaches genome-wide significance supporting it as a true diabetes locus in spite of only nominally significant replication in our DIREVA study. Subdividing cases by suptype could thus be a powerful strategy for identification of new diabetes loci with the additional benefit of information about pathways affected by the loci and the mechanisms underlying each subtype.

While the ANDIS participants were all recruited within one year from diabetes diagnosis, DIREVA also included individuals with longer duration of disease. The fact that we can replicate so many of the genetic findings from ANDIS supports that clusters identified in patients with longer duration are largely similar to clusters from parameters measured at diabetes onset, but the smaller effect sizes and less distinct associations suggest that there could be some misclassification. This is in line with a small study in the German Diabetes Study where 27% of individuals changed subtype if clustering was repeated after 5 years.^2^ A recent study by Bello-Chavolla et al, used an alternative classification approach in cohorts of Mexican origin. The same diabetes subtypes were identified using self-normalizing neural networks and surrogate classification.^4^ In this study a large proportion of SIDD patients transferred to MOD and MARD when clustering was repeated after up to 2 years, whereas other subtypes were more stable. This indicates that the classification method might need to be adapted for optimal classification of long duration patients.

Power is a weakness for the analysis of single loci. Future larger studies should combine clustering of T2D with genetic studies to identify more predictive SNPs and possibly also genetic variants predicting treatment response in a subgroup.

In conclusion, the data provide strong evidence for genetically different backgrounds of the subtypes. Especially SIRD stands out with seemingly a largely beta-cell independent pathogenesis. This study thereby shows that the new subtypes are partially distinct and that subclassification of T2D is valuable for pre-clinical research and important clinically, opening up for better targeted intervention studies.

## Methods

### Study populations

The ANDIS (All New Diabetics in Scania) project (http://andis.ludc.med.lu.se/) aims to recruit all incident cases of diabetes within Scania (Skåne) County in southern Sweden, which has about 1,200,000 inhabitants. All health care providers in the region were invited; the current registration covered the period January 1st 2008 until November 2016 during which 177 clinics registered 14,625 patients aged 0-96 years, within a median of 40 days (IQR 12-99) after diagnosis. Individuals were assigned to clusters/subtypes as previously described.^1^ Patient characteristics are presented in Table S1.

The Malmö Diet and Cancer (MDC) Study includes individuals from Malmö, Sweden, born between 1923 and 1950. Diabetes-free individuals (n=2744) from the MDC cardiovascular arm (MDC-CVA) re-examination cohort (age 61-85) were used as controls in genetic analyses.^21^ DIREVA (Diabetes Registry Vaasa) from Western Finland (∼170,000 inhabitants) includes 5107 individuals with diabetes recruited 2009-2014 in the Vaasa hospital district. Individuals were assigned to clusters/subtypes as previously described.^1^

The Botnia Study has recruited patients with T2D and their family members in the area of five primary health care centers in Western Finland since 1990.^22^ Unrelated (based on estimated genetic relationships) diabetes free individuals were used as controls for DIREVA (Table S1).

### Genotyping

ANDIS and DIREVA were genotyped with InfiniumCoreExome-24v1-1 BeadChip arrays (Illumina, San Diego, CA, USA), at Lund University Diabetes Centre, Malmö, Sweden. MDC was genotyped at the Broad genotyping facility using Infinium OmniExpressExome-8 version 1.0 BeadChip arrays (Illumina, San Diego, CA, USA). Botnia controls were genotyped using Illumina Global Screening array-24v1 at Regeneron Pharmaceuticals Inc, NY, US.

For each of the genetic datasets, standard quality control was performed.^23^ Samples were excluded for ambiguous gender, call rate < 95%, and any duplicate or related individuals (pi_hat ≥ 0.2). SNP were excluded for monomorphic SNPs, SNPs with MAF < 0.05, and SNPs with missingness rate > 0.05.

Genotypes from the ANDIS (12770 individuals) and MDC (3344 individuals) cohorts were merged using PLINK,^24^ including only SNPs present on both genotyping arrays. After the merge, 16804 individuals and 324063 SNPs passed QC.

Genotype imputation was performed at the HRC Michigan Server^25^ for autosomal chromosomes.

### Family history

Family history of diabetes (FHD) was self-reported at registration in ANDIS. In the ESTRID^5^ sub-study of ANDIS, conducted 2010-2019, all individuals with LADA and a random sample of individuals with T2D and density matched randomly sampled diabetes free controls (≥35 years, n=2290) from the same region completed a detailed questionnaire on FHD, lifestyle and health.

FHD was reported in first (mother, father, sisters, brothers) and second-degree relatives (grandparents, aunts, uncles). FHD–T1D was defined as a relative with diagnosis at age < 40 years and insulin treatment and, if otherwise, as FHD–T2D.

Conditional logistic regression was used for the association between FHD and subtype, adjusting for sex. Differences in proportions were calculated by χ2.

### Genetic risks cores analysis (GRS)

T1D and T2D polygenic risk scores were constructed using r^2^=0.1, and 250kb window, as implemented in PLINK v1.9/2. The T2D weighted genetic risk scores (T2D-wGRS) for glycemic traits were calculated based on the genetic effect of only known T2D associated variants (n=353, MAF>1% in ANDIS) or their proxies (r2 >0.8) after clumping (r^2^ =0.5, 250kb window) in PLINK. GRSs for adiposity related traits and lipids, were calculated using only genome-wide significant SNPs retained after clumping (r^2^=0.1, and 250kb window) for each trait. Association analysis with diabetes subtypes was done by logistic regression of inversely normally transformed GRS adjusting for the first 10 principal components (PCs) in R.

### Genome-wide association analysis

GWAS analysis was done in SNPTEST (v2.5.2)^26^ using frequentist association analysis score method adjusting for sex and first 10 PCs. Individuals of non-European origin (1240 individuals) and first-second degree relatives (995 individuals) were excluded from analysis. Results were filtered based on minor allele frequency (MAF > 0.05), Hardy-Weinberg (HWE > 5e^-07^) and imputation info scores (INFO > 0.4). Manhattan plots and QQplots were generated in R using QQMAN package.^27^

## Data Availability

The data that support the findings of this study are available upon request but restrictions apply. Individual level data are not publicly available due to ethical and legal restrictions related to the Swedish Biobanks in Medical Care Act (2002:297) and the Personal Data Act (1998:204), EU's General Data Protection Regulation (GDPR) 2016/679, and the Data Protection Act 2018:218.

## Author contributions

DMA, OPD, RBP, JF, MM, TT, LG and EA contributed to the conception of the work. AK, RGC, JB, OM, SC, OH, TT, LG and EA contributed to the data collection. DMA, OPD, RH, MÅ, AM, MU and EA contributed to the data analysis. DMA, OPD, TT, LG and EA drafted the article. All authors contributed to the interpretation of data and critical revision of the Article. All authors gave final approval of the version to be published.

## Funding

This study was supported by grants from the Swedish Research Council (project grant 521-2010-3490 and infrastructure grants 2010-5983, 2012-5538, and 2014-6395 to LG; project grant 2017-02688 to EA; Linnaeus grant 349-2006-237; and a strategic research grant 2009-1039 to LG), a European Research Council Advanced Research grant (GA 269045), a Vinnova Swelife grant, and grants from the Academy of Finland (263401and 267882 to LG), Sigrid Juselius Foundation, Novo Nordisk Foundation (NNF18OC0034408 to EA), Scania University Hospital (ALF grant), Diabetes Wellness Sweden (25-420 PG), the Swedish Heart and Lung Foundation, the Swedish Diabetes Foundation, the Crafoord Foundation, the Albert Påhlsson Foundation. This project was also financially supported by the Swedish Foundation for Strategic Research (IRC15-0067). The genotyping of ANDIS was funded by Pfizer Inc. Cambridge, MA, US. DIREVA was supported by the Vasa Hospital district, Jakobstadsnejden Heart Foundation, Folkhalsan Research Foundation, and Ollqvist Foundation (to TT and AK). The Botnia Study (L.G., T.T.) have been financially supported by grants from Folkhälsan Research Foundation, the Sigrid Juselius Foundation, The Academy of Finland (grants no. 263401, 267882, 312063 to LG, 312072 to TT), University of Helsinki, Nordic Center of Excellence in Disease Genetics, EU (EXGENESIS, MOSAIC FP7-600914), Ollqvist Foundation, Swedish Cultural Foundation in Finland, Finnish Diabetes Research Foundation, Foundation for Life and Health in Finland, Signe and Ane Gyllenberg Foundation, Finnish Medical Society, Paavo Nurmi Foundation, State Research Funding via the Helsinki University Hospital, Perklén Foundation, Närpes Health Care Foundation and Ahokas Foundation. The study has also been supported by the Ministry of Education in Finland, Municipal Heath Care Center and Hospital in Jakobstad and Health Care Centers in Vasa, Närpes and Korsholm. The research leading to these results has received funding from the European Research Council under the European Union’s Seventh Framework Programme (FP7/2007-2013) / ERC grant agreement n° 269045. The skillful assistance of the Botnia Study Group is gratefully acknowledged. The genotyping of Botnia was funded by Regeneron Pharmaceuticals Inc. NY, US.

We thank all patients and health-care providers for their support and willingness to participate. We also thank Johan Hultman, Mattias Borell, Jasmina Kravic, Gabriella Gremsperger, Maria Sterner, Malin Neptin and Ulrika Blom-Nilsson for excellent technical and administrative support; Region Skåne (Scania County); and the ANDIS steering committee for their support.

## Supplementary Figures

**Figure S1.**
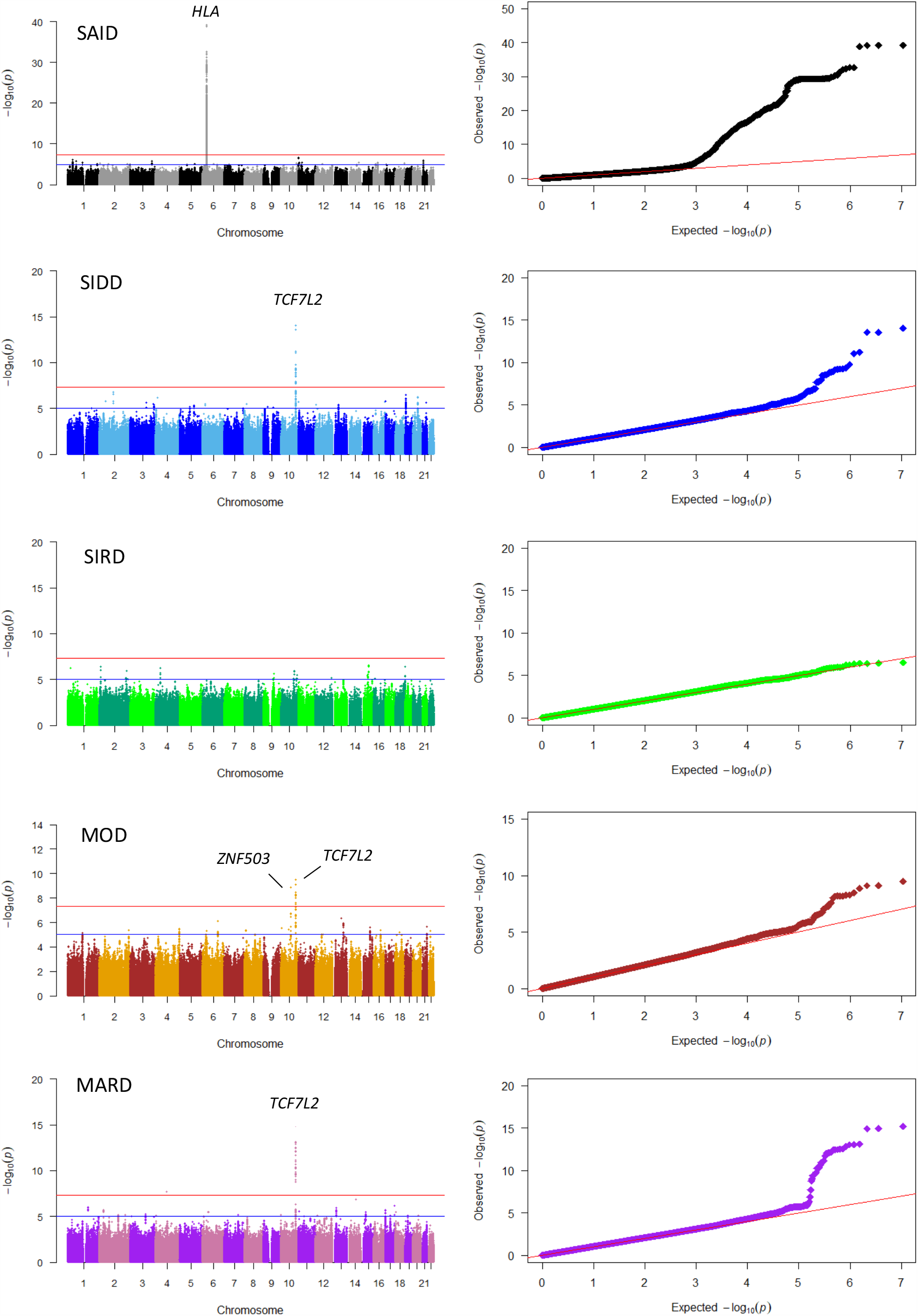
Manhattan and QQ plots of GWAS in ANDIS-MDC. Variants in the *HLA-DQB1* locus were associated with SAID (rs9273368 OR=2.893[2.314-3.100], p=6.50⨯10^−40^). The rs7903146 *TCF7L2* locus was associated with SIDD (OR=1.559[1.394-1.745], p=8.59⨯10^−15^), MOD (OR=1.402[1.262-1.557], P=3.13⨯10^−10^) and MARD (1.417[1.302-1.542], p=6.07⨯10^−16^). Rs10824307 near the *ZNF503* gene was associated with MOD (OR=1.347 (1.219-1.487), p=1.3⨯10^−9^.

## Regeneron Genetics Center Banner Author List and Contribution Statements

All authors/contributors are listed in alphabetical order.

### RGC Management and Leadership Team

Goncalo Abecasis, Ph.D., Aris Baras, M.D., Michael Cantor, M.D., Giovanni Coppola, M.D., Aris Economides, Ph.D., Luca A. Lotta, M.D., Ph.D., John D. Overton, Ph.D., Jeffrey G. Reid, Ph.D., Alan Shuldiner, M.D.

Contribution: All authors contributed to securing funding, study design and oversight. All authors reviewed the final version of the manuscript.

### Sequencing and Lab Operations

Christina Beechert, Caitlin Forsythe, M.S., Erin D. Fuller, Zhenhua Gu, M.S., Michael Lattari, Alexander Lopez, M.S., John D. Overton, Ph.D., Thomas D. Schleicher, M.S., Maria Sotiropoulos Padilla, M.S., Louis Widom, Sarah E. Wolf, M.S., Manasi Pradhan, M.S., Kia Manoochehri, Ricardo H. Ulloa.

Contribution: C.B., C.F., A.L., and J.D.O. performed and are responsible for sample genotyping. C.B, C.F., E.D.F., M.L., M.S.P., L.W., S.E.W., A.L., and J.D.O. performed and are responsible for exome sequencing. T.D.S., Z.G., A.L., and J.D.O. conceived and are responsible for laboratory automation. M.P., K.M., R.U., and J.D.O are responsible for sample tracking and the library information management system.

### Genome Informatics

Xiaodong Bai, Ph.D., Suganthi Balasubramanian, Ph.D., Andrew Blumenfeld, Gisu Eom, Lukas Habegger, Ph.D., Alicia Hawes, B.S., Shareef Khalid, Jeffrey G. Reid, Ph.D., Evan K. Maxwell, Ph.D., William Salerno, Ph.D., Jeffrey C. Staples, Ph.D.

Contribution: X.B., A.H., W.S. and J.G.R. performed and are responsible for analysis needed to produce exome and genotype data. G.E. and J.G.R. provided compute infrastructure development and operational support. S.K., S.B., and J.G.R. provide variant and gene annotations and their functional interpretation of variants. E.M., J.S., A.B., L.H., J.G.R. conceived and are responsible for creating, developing, and deploying analysis platforms and computational methods for analyzing genomic data.

### Research Program Management

Marcus B. Jones, Ph.D., Lyndon J. Mitnaul, Ph.D.

Contribution: All authors contributed to the management and coordination of all research activities, planning and execution. All authors contributed to the review process for the final version of the manuscript.

